# Validation of an extraction-free RT-PCR protocol for detection of SARS-CoV2 RNA

**DOI:** 10.1101/2020.04.29.20085910

**Authors:** Julianne R Brown, Laura Atkinson, Divya Shah, Kathryn Harris

## Abstract

In light of supply chain failures for reagents and consumables needed for purification of nucleic acid for detection of SARS-CoV-2 RNA by RT-PCR, we aim to verify the performance and utility of a non-extraction protocol for RT-PCR (“direct RT-PCR”).

We report improved sensitivity compared to earlier reports of direct RT-PCR testing of swab samples, in particular at the lower limit of detection (sensitivity 93% overall; 100% for specimens with high to moderate viral titre, Ct <34; 81% for specimens with a low viral titre, Ct ≥34). Sensitivity is improved (from 90 to 93%) by testing in duplicate. We recommend swabs are re-suspended in water to minimise PCR inhibition. A cellular target is necessary to control for PCR inhibition and specimen quality.

Direct RT-PCR is best suited to population level screening where results are not clinically actionable, however in the event of a critical supply chain failure direct RT-PCR is fit for purpose for the detection of SARS-CoV-2 infection.

The results from our study offer front-line laboratories additional reagent options for performing extraction-free RT-PCR protocols.

## INTRODUCTION

The 2019/2020 SARS-CoV-2 global pandemic has seen the need for large-scale RT-PCR testing. The gold standard for detection of viruses causing infection is purification/extraction of nucleic acid followed by one-step real-time RT-PCR. However testing efforts have been hampered by global shortages of critical laboratory reagents, in particular reagents and consumables for RNA purification.

In light of the global demand for testing and the bottleneck in capacity caused by nucleic acid extraction, clinical laboratories have considered turning to real-time RT-PCR direct from clinical specimens without prior nucleic acid extraction, herein referred to as direct RT-PCR.

Initial reports for direct RT-PCR are promising, with 92% (Bruce et al., 2020) and 98% (Grant et al., 2020) reported sensitivity. However there is an estimated 10-fold increase in the lower-limit of detection compared to RT-PCR from purified RNA (Grant et al., 2020) and a maximum specimen input volume of 2-3 μl due to the presence of inhibitory substances (Grant et al., 2020, Bruce et al., 2020).

We aim to verify the performance and utility of direct RT-PCR in clinical practice. In particular we test direct RT-PCR using different PCR mastermixes to those reported previously with the aim of improving sensitivity and lower limit of detection and potentially recommend alternative mastermixes in which direct RT-PCR is possible.

## METHODS

### Test material

#### Specimens

All specimens were nose or throat swabs submitted to our diagnostic laboratory for detection of SARS-CoV-2 RNA. Dry swabs were re-suspended with 600 or 1,200 μl nuclease-free water (for single or double swabs, respectively).

Total nucleic acid was purified from 200 μl swab suspension fluid using the Hamilton Nimbus (with Kingfisher Presto) and Omega Biotek Mag-Bind Viral DNA/RNA kit with 100 μl elution volume. RT-PCR was performed as described below using the purified nucleic acid.

Direct PCR was performed using residual swab suspension fluid either on the same day as nucleic acid purification or after storage at −80°C. PCR set-up was performed in a biosafety cabinet for samples that had not been heat inactivated.

#### Mastermixes

Ten specimens known to be SARS-CoV-2 RT-PCR positive from purified nucleic acid (Ct from purified nucleic acid range 22-39) were tested in triplicate by direct RT-PCR using a 2 μl sample input in 25 μl reactions with the following mastermixes; One Step PrimeScript™ III RT-PCR Kit mastermix [Takara], Quantifast Multiplex RT-PCR +R mastermix [Qiagen] and TaqPath 1-Step RT-qPCR MasterMix {Thermo Fisher], These mastermixes were chosen due to their previously reported performance in SARS-CoV-2 PCR (Brown et al., 2020). Cycling conditions were as described previously (Brown et al., 2020).

#### Specimen input volume

Ten specimens known to be SARS-CoV-2 RT-PCR positive from purified nucleic acid (Ct range from purified nucleic acid 22-39) were tested in triplicate by direct RT-PCR using One Step PrimeScript™ III RT-PCR Kit mastermix [Takara] with a specimen input volume of 2, 5 and 10 μl.

#### Polymerase Chain Reaction

We performed real-time RT-PCR targeting the SARS-CoV-2 N gene (Grant et al., 2020) as follows. Each 25 μl reaction consisted of 12.5 μl One Step PrimeScript™ III RT-PCR Kit mastermix [Takara], 0.5 μl Rox Dye II, 0.4 μM NgeneTaq-forward primer, 0.6μM NgeneTaq-reverse primer, 0.3 μM NgeneTaq-probe, 10 μl swab fluid or 7.5 μl purified nucleic acid and nuclease-free water.

Where specified, RT-PCR targeting the cellular human target RNAseP was performed as described above using primers described previously (Emery et al., 2004), with 0.5 μM of each primer and 0.125 μM probe per reaction.

All PCR reactions were run on a Quantstudio 5 thermocycler [Thermo Fisher],

#### Takara mastermix comparison

Twenty-one SARS-CoV-2 positive specimens (median Ct from purified nucleic acid 31, range 20–40) were tested, in duplicate, by direct RT-PCR with a 10 μl input volume and One Step PrimeScript™ III RT-PCR Kit [Takara] or PrimeDirect Probe RT-qPCR Mix [Takara]. PrimeDirect mastermix was chosen as it is marketed specifically for use in direct RT-PCR. Specimens were tested by direct RT-PCR without prior heat treatment.

Based on manufacturer recommendations, specimens tested with PrimeDirect were also heated for 10 minutes at 99°C prior to direct RT-PCR. Pre-heated and un-heated PrimeDirect reactions underwent RT-PCR with different cycling conditions. Briefly, cycling conditions for unheated samples consisted of 90 °C for 3 minutes, followed by 60 °C for 5 minutes and 45 cycles of 95 °C for 5 seconds and 60 °C for 30 seconds. Cycling conditions for heated samples consisted of 95 °C for 10 seconds, followed by 60 °C for 5 minutes and 45 cycles of 95 °C for 5 seconds and 60 °C for 30 seconds.

#### Heat lysis prior to direct RT-PCR

Seventy-seven SARS-CoV-2 positive specimens (median Ct from purified nucleic acid 31, range 20-42) were tested by direct RT-PCR with 10 μl input volume and with PrimeScript mastermix [Takara], without any prior heat treatment. A subset of these, 53/77, were additionally heated at 56 °C for 15 minutes to lyse epithelial cells and inactivate SARS-CoV2 prior to direct RT-PCR.

#### Viral transport media

Nineteen fresh (not stored at −80 °C) swabs in viral transport media were tested in duplicate by direct RT-PCR with a 10 μl input and PrimeScript [Takara] mastermix. Four of the nineteen (4/19) swabs were known to be SARS-CoV-2 positive (Ct 25, 34, 37 and 40) and 15/19 were SARS-CoV-2 negative by PCR from purified nucleic acid. All (19/19) were tested by direct-RT PCR targeting SARS-CoV-2 RNA and the human target RNAseP.

An additional 9 dry swabs, resuspended in nuclease free water, were tested by direct RT-PCR targeting RNAseP, for comparison to the VTM swabs.

## RESULTS

### Mastermixes

Of the ten specimens tested by direct RT-PCR with a 2 μl input volume, 3/10 were PCR positive in at least one of the three replicates with Qiagen Quantifast mastermix, 8/10 with Thermo Fisher TaqPath mastermix and 9/10 with Takara PrimeScript mastermix.

Zero (0/10) were PCR positive across all three replicates with Qiagen Quantifast mastermix, 7/10 with Thermo Fisher TaqPath mastermix and 7/10 with Takara PrimeScript mastermix (Table 1).

**Table 1.**
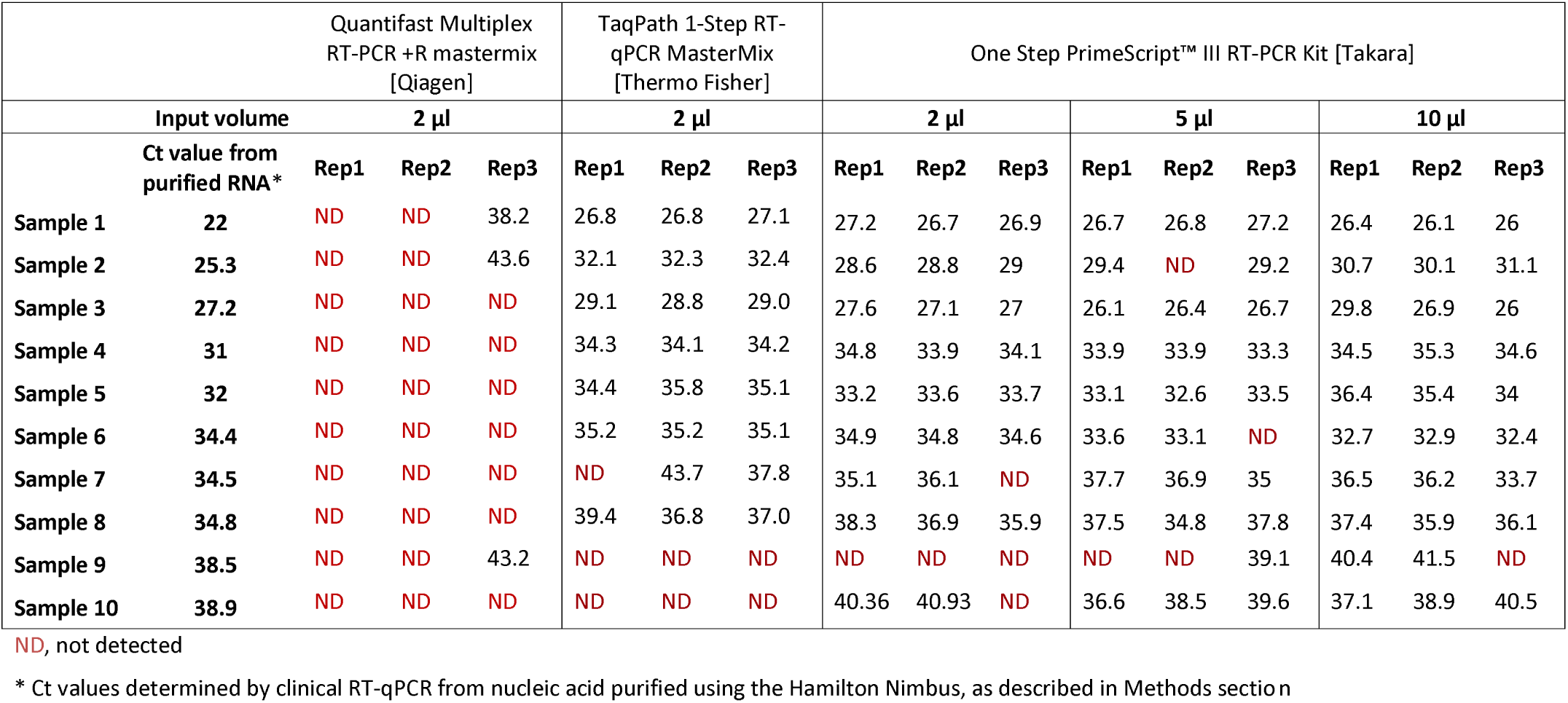
Ct values obtained by direct RT-PCR for 10 SARS-CoV-2 positive samples with multiple mastermixes and sample input volumes

|  |  | Quantifast Multiplex<br>RT-PCR +R mastermix<br>[Qiagen] |  |  | TaqPath 1-Step RT-qPCR MasterMix<br>[Thermo Fisher] |  |  | One Step PrimeScript™ III RT-PCR Kit [Takara] |  |  |  |  |  |  |  |  |
| --- | --- | --- | --- | --- | --- | --- | --- | --- | --- | --- | --- | --- | --- | --- | --- | --- |
| Input volume |  | 2 µl |  |  | 2 µl |  |  | 2 µl |  |  | 5 µl |  |  | 10 µl |  |  |
|  | Ct value from<br>purified RNA* | Rep1 | Rep2 | Rep3 | Rep1 | Rep2 | Rep3 | Rep1 | Rep2 | Rep3 | Rep1 | Rep2 | Rep3 | Rep1 | Rep2 | Rep3 |
| Sample 1 | 22 | ND | ND | 38.2 | 26.8 | 26.8 | 27.1 | 27.2 | 26.7 | 26.9 | 26.7 | 26.8 | 27.2 | 26.4 | 26.1 | 26 |
| Sample 2 | 25.3 | ND | ND | 43.6 | 32.1 | 32.3 | 32.4 | 28.6 | 28.8 | 29 | 29.4 | ND | 29.2 | 30.7 | 30.1 | 31.1 |
| Sample 3 | 27.2 | ND | ND | ND | 29.1 | 28.8 | 29.0 | 27.6 | 27.1 | 27 | 26.1 | 26.4 | 26.7 | 29.8 | 26.9 | 26 |
| Sample 4 | 31 | ND | ND | ND | 34.3 | 34.1 | 34.2 | 34.8 | 33.9 | 34.1 | 33.9 | 33.9 | 33.3 | 34.5 | 35.3 | 34.6 |
| Sample 5 | 32 | ND | ND | ND | 34.4 | 35.8 | 35.1 | 33.2 | 33.6 | 33.7 | 33.1 | 32.6 | 33.5 | 36.4 | 35.4 | 34 |
| Sample 6 | 34.4 | ND | ND | ND | 35.2 | 35.2 | 35.1 | 34.9 | 34.8 | 34.6 | 33.6 | 33.1 | ND | 32.7 | 32.9 | 32.4 |
| Sample 7 | 34.5 | ND | ND | ND | ND | 43.7 | 37.8 | 35.1 | 36.1 | ND | 37.7 | 36.9 | 35 | 36.5 | 36.2 | 33.7 |
| Sample 8 | 34.8 | ND | ND | ND | 39.4 | 36.8 | 37.0 | 38.3 | 36.9 | 35.9 | 37.5 | 34.8 | 37.8 | 37.4 | 35.9 | 36.1 |
| Sample 9 | 38.5 | ND | ND | 43.2 | ND | ND | ND | ND | ND | ND | ND | ND | 39.1 | 40.4 | 41.5 | ND |
| Sample 10 | 38.9 | ND | ND | ND | ND | ND | ND | 40.36 | 40.93 | ND | 36.6 | 38.5 | 39.6 | 37.1 | 38.9 | 40.5 |

ND, not detected
\* Ct values determined by clinical RT-qPCR from nucleic acid purified using the Hamilton Nimbus, as described in Methods section

One Step PrimeScript™ III RT-PCR Kit [Takara] was chosen for direct RT-PCR going forward.

### Specimen Input Volume

Of the ten specimens tested by direct RT-PCR with variable input volumes, 7/10 were PCR positive across all three replicates with a 2μl input volume, 7/10 with a 5 μl input volume and 9/10 with a 10 μl input volume (Table 1).

A sample input volume of 10μl was chosen for direct RT-PCR going forwards.

### Takara mastermix comparison

In our hands the overall sensitivity of PrimeDirect Probe RT-qPCR Mix [Takara] is 57-62%, compared to 95% for One Step PrimeScript™ III RT-PCR Kit [Takara] (Table 2).

**Table 2.** Detection sensitivity of direct RT-PCR in Takara PrimeScript and PrimeDirect mastermixes. All samples (n = 21) were tested in duplicate

|  | Detected in 2/2 duplicates |  |  | Detected in at least 1/2 duplicates |  |  |
| --- | --- | --- | --- | --- | --- | --- |
| **SARS-CoV-2 RNA titre\*** | **PrimeScript (unheated)** | **PrimeDirect (unheated)** | **PrimeDirect (heated 99 °C)** | **PrimeScript (unheated)** | **PrimeDirect (unheated)** | **PrimeDirect (heated 99 °C)** |
| **High (Ct <25)** | 4/4<br>(100%) | 4/4<br>(100%) | 4/4<br>(100%) | 4/4<br>(100%) | 4/4<br>(100%) | 4/4<br>(100%) |
| **Intermediate (Ct 25–33)** | 12/12<br>(100%) | 3/12<br>(25%) | 7/12<br>(58.3%) | 12/12<br>(100%) | 8/12<br>(66.7%) | 9/12<br>(75%) |
| **Low (Ct ≥34)** | 2/5<br>(40%) | 0/5<br>(0%) | 0/5<br>(0%) | 4/5<br>(80%) | 0/5<br>(0%) | 0/5<br>(0%) |
| **TOTAL** | 18/21<br>**(85.7%)** | 7/21<br>**(33.3%)** | 11/21<br>**(52.4%)** | 20/21<br>**(95.2%)** | 12/21<br>**(57.1%)** | 13/21<br>**(61.9%)** |

\* Ct values determined by clinical RT-qPCR from nucleic acid purified using the Hamilton Nimbus, as described in Methods section

One Step PrimeScript™ III RT-PCR Kit [Takara] was used for direct RT-PCR going forward.

### Heat lysis prior to direct PCR

Overall sensitivity was improved when specimens were not heat –treated prior to direct RT-PCR (90-93% sensitivity) compared to when specimens were heated to 56 °C for 15 minutes prior to direct RT-PCR (81–90% sensitivity) (Table 3).

**Table 3.** Detection sensitivity of direct RT-PCR in PrimeScript mastermix [Takara] without prior heating (n=77) and with prior heat-treatment at 56 °C for 15 minutes (n=53). The heat-treated specimens are a subset of the unheated specimens. All samples (n = 77) were tested in duplicate. One sample was excluded from the results as both SARS-CoV-2 and RNaseP were not detected therefore would be reported as an invalid result.

|  | Detected in 2/2 duplicates |  | Detected in at least 1/2 duplicates |  |
| --- | --- | --- | --- | --- |
| **SARS-CoV-2 RNA titre\*** | **Unheated** | **Heated 56 °C** | **Unheated** | **Heated 56 °C** |
| **High (Ct <25)** | 12/12<br>(100%) | 7/7<br>(100%) | 12/12<br>(100%) | 7/7<br>(100%) |
| **Intermediate (Ct 25–33)** | 38/38<br>(100%) | 24/25<br>(96%) | 38/38<br>(100%) | 24/25<br>(96%) |
| **Low (Ct ≥34)** | 18/26<br>(69%) | 11/20<br>(55%) | 21/26<br>(81%) | 16/20<br>(80%) |
| **TOTAL** | 68/76<br>**(90%)** | 42/52<br>**(81%)** | 71/76<br>**(93%)** | 47/52<br>**(90%)** |

\* Ct values determined by clinical RT-qPCR from nucleic acid purified using the Hamilton Nimbus, as described in Methods section

Sensitivity for specimens with a Ct value <34 from purified nucleic acid was 100%, but reduced to 69–81% and 55–80% for unheated and heated samples with a Ct value ≥34 from purified nucleic acid, respectively (Table 3).

Specimens were not pre-heated for direct RT-PCR going forwards.

### Viral transport media and RNAseP

Seven (7/19, 36.8%) of swabs in viral transport media (VTM) were positive in both duplicates for the human target RNAseP; 15/19 (78.9%) were RNAseP positive in at least one (1/2) of the duplicates. None of the SARS-CoV-2 positive VTM swabs (0/4, 0%) were detected by direct-RT PCR, including the swab which was positive with Ct 25 by PCR from purified nucleic acid; this swab was RNAseP PCR positive (Ct 25-26) by direct RT-PCR.

RNaseP was detected by direct RT-PCR in all (9/9, 100%) dry swabs resuspended in water, in both duplicates. RNAse P was detected in all (90/90,100%) of specimens from purified nucleic acid.

The median RNAseP Ct value in dry swabs with water was 26 (range 23-30) and 27 in RNAseP positive VTM swabs (range 23-31). The median RNAseP Ct value from purified nucleic acid (n=90) was 27 (range 23-35).

### Change in PCR Ct value

The median change in Ct value between samples tested by RT-PCR from purified nucleic acid and by direct RT-PCR is +1.9 cycles (range −2.7 to +10.1) (Table 4). There is a greater change in Ct values (median +2.4 cycles) for high titre specimens (Ct <25).

**Table 4.** Change in Ct value between direct RT-PCR and RT-PCR from purified nucleic acid. Calculated from 139 direct RT-PCR positive results, from 76 swabs.

| **SARS-CoV-2 RNA titre\*** | **Median Ct value change** | **Range of Ct value change** |
| --- | --- | --- |
| **High (Ct <25)** | +2.4 | +0.1 – 8.3 |
| **Intermediate (Ct 25–33)** | +1.9 | -2.7 – 10.1 |
| **Low (Ct ≥34)** | +1.6 | -2.6 – 6.7 |
| **TOTAL** | **+1.9** | **-2.7 – +10.1** |

\* Ct values determined by clinical RT-qPCR from nucleic acid purified using the Hamilton Nimbus, as described in Methods section

### Estimated detection rates

Since 23^rd^ March 2020 our laboratory has processed 1698 swabs for detection of SARS-CoV-2 RNA, of which 575/1698 were SARS-CoV-2 PCR positive from purified nucleic acid.

Based on the data presented here, we estimate that 8-13% (46-75/575) of these would have been missed if tested using direct RT-PCR with 10 μl of sample with One Step PrimeScript™ III RT-PCR Kit [Takara], all with an original Ct value ≥34 (Figure 1, Table 5). The range in estimated false negatives is dependent on whether specimens are tested singularly or in duplicate.

**Figure 1.**
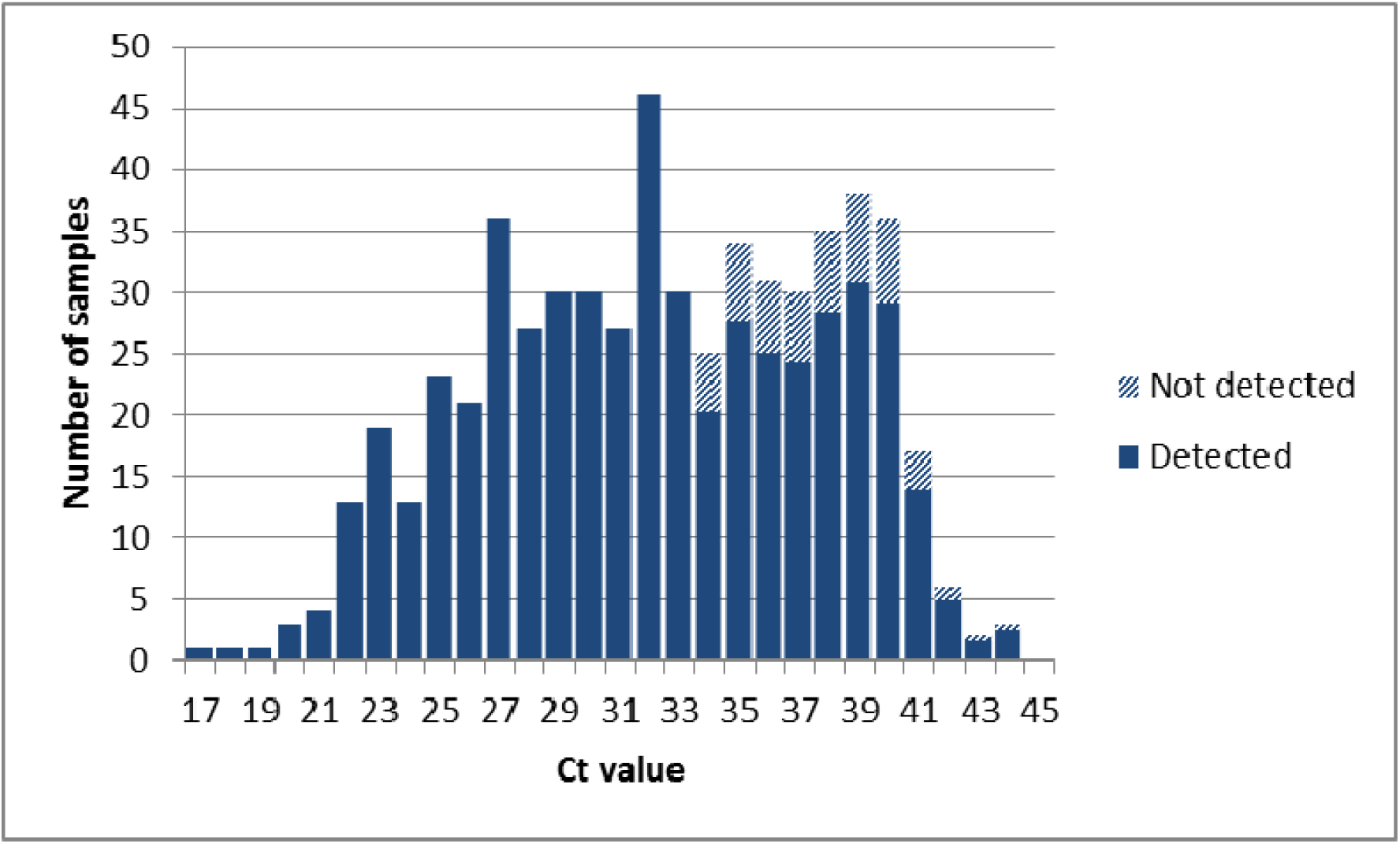
Distribution of Ct values from SARS-CoV-2 positive swabs (n = 575/1698) tested by RT-PCR from purified nucleic acid in our laboratory from 23^rd^ March to 23^rd^ April 2020. Results are divided into those that we would expect to have detected (529/575) and not detected (46/575) using the extraction-free protocol described in this study (based on specimens tested in duplicate with 81% sensitivity for specimens with Ct ≥34).

## CONCLUSIONS

### Variability in Mastermix

Given that direct-RT PCR is highly variable between mastermixes, each laboratory must perform their own validation using the mastermixes they have available. Our study provides an additional mastermix option to those reported previously, namely One Step PrimeScript™ III RT-PCR Kit [Takara]. TaqPath 1-Step RT-qPCR MasterMix [Thermo Fisher] may also be suitable for direct RT-PCR, however it was not explored further in this study due to current lack of availability.

PrimeDirect Probe RT-qPCR Mix [Takara], which is marketed specifically for use in direct RT-PCR, and Quantifast Multiplex RT-PCR +R mastermix [Qiagen] were not fit for purpose.

One Step PrimeScript™ III RT-PCR Kit mastermix [Takara] allows a 10 μl specimen input volume per reaction, compared to 2-3 μl with the mastermixes from previous reports (Grant et al., 2020, Bruce et al., 2020), which we suggest as the reason for the improved performance for low-level positives. Moreover a greater input volume is less likely to be susceptible to operator error; especially when the PCR set-up is conducted in a biosafety cabinet with limited visibility compared to the open bench.

### Effect of swab re-suspension media

Swabs suspended in viral transport media were found to be inhibitory to direct-RT PCR. We recommend that for the purpose of direct RT-PCR specimens should be collected as dry flocked swabs and resuspended in nuclease-free water.

The suitability of direct RT-PCR for other specimen types, for example nasopharyngeal aspirates (NPAs), remains to be determined. However we suggest it would be prudent that, where possible, specimens such as NPAs are collected using water instead of saline to minimise the risk of PCR inhibition.

### Sensitivity

In our hands, using PrimeScript mastermix [Takara] and 10 μl input volume, direct RT-PCR to detect SARS-CoV-2 RNA from dry swabs resuspended in water had a sensitivity of up to 93%, if samples are tested in duplicate. This is less than the 98% reported by Grant *et al*. (Grant et al., 2020) however Grant *et al*. did not report sensitivity in specimens with a Ct value greater than 32 which may have caused an over-estimation of their reported sensitivity. In our study we detected 100% (45/45) of specimens with a Ct value ≤32.

Our overall sensitivity of up to 93% is comparable to that reported by Bruce *et al*. (Bruce et al., 2020). However Bruce *et al*. tested a smaller proportion of low positive samples (17/150 specimens, 11%, Ct >30) than our study (42/76 specimens, 55%, Ct >30 and 26/76 specimens, 34%, Ct ≥34). Bruce *et al* reported a sensitivity of 35% in specimens with a Ct >30, compared to in our study a sensitivity of 81-88% in specimens with Ct >30 (34/42 SARS-CoV-2 positive in 2/2 duplicates and 37/42 positive in at least 1/2 duplicates, respectively). Consequently the sensitivity for low level positives appears to be improved with the experimental conditions reported in this study, however this cannot be confirmed without a side-by-side comparison using the same specimens.

We estimate that of the 1698 samples tested in our service to date with nucleic acid extraction, 8-13% of positives (46-75/575; 2.7% of all specimens) would have been missed by direct RT-PCR. The testing period during which these specimens were collected included a period of clearance screening for essential healthcare workers recovering from confirmed SARS-CoV-2 infection. These clearance screens often presented with low level positives (data not shown), therefore our data may be skewed towards low level positives. Testing programmes that do not include clearance screening may have a lower proportion of low positive specimens, and therefore potentially improved overall sensitivity.

Given that sensitivity of direct RT-PCR is less than 100%, it is likely that some SARS-CoV-2 positive specimens will be missed using direct RT-PCR, especially those with a low viral load. To maximise detection of SARS-CoV-2 RNA with an extraction-free protocol it is critical that laboratories implement an optimised combination of PCR primers and mastermix, such as those we have previously reported (Brown et al., 2020), to maximise sensitivity. Testing specimens in duplicate will help to improve the sensitivity of direct RT-PCR, as demonstrated in our study in which sensitivity was improved from 90 to 93% by testing specimens in duplicate.

### Shift in Ct values

We report a shift in Ct values when samples are tested by direct RT-PCR, compared to RT-PCR from purified nucleic acid, with positives detected up to 10 cycles later but on average 2 cycles later. Having said this, the shift in Ct values is less pronounced at the lower end of detection (median change in Ct +1.6) and therefore does not have much of an effect on overall sensitivity, contrary to what might have been expected.

Consequently we suggest that inferences are not made from Ct values, with results treated as qualitative positive or negative. Furthermore, given the loss in sensitivity seen with direct RT-PCR, very low positive results (Ct ≥38) should be reported as positive, with no comment on it being a low positive or equivocal result.

### Clinical utility

In the event of a catastrophic supply chain failure for nucleic acid extraction reagents or consumables, direct RT-PCR using the non-extraction protocol described in this study, with overall sensitivity of 93%, would be an adequate option for detection of SARS-CoV-2 RNA. To maximise sensitivity and minimise false-negative results we recommend testing samples in duplicate. Inclusion of a cellular target, such as RNAseP, in multiplex with SARS-CoV-2 is essential to control for PCR inhibition and specimen quality. This is particularly important for samples collected via home swabbing programmes where the quality of the sample may be reduced.

We suggest that direct RT-PCR is particularly suited to population-level screening for SARS-CoV-2, where the results are not clinically actionable. In this setting an extraction-free protocol unlocks the potential for high through-put and rapid testing that is robust in the face of supply chain challenges for nucleic acid extraction reagents and consumables.

Extraction-free protocols are not suited to scenarios where one expects to see a high proportion of low level positives, such as return-to-work clearance screening of individuals recovering from confirmed SARS-CoV-2 infection.

**Table 5.** Estimated false negative rate for all samples tested to date in our laboratory (n=1698), if they were to be tested by direct RT-PCR with the non-extraction protocol described in this study

| **SARS-CoV-2 RNA titre\*** | **Number of samples SARS-CoV-2 positive from purified RNA \*** | **Estimated detection rate by direct RT-PCR†** | **Estimated false-negatives** |
| --- | --- | --- | --- |
| **High (Ct <25)** | 78 | 100% | 0 |
| **Intermediate (Ct 25–33)** | 255 | 100% | 0 |
| **Low (Ct ≥34)** | 242 | 69–81%** | 46–75 specimens |
| **TOTAL** | 575/1698 | 90–93% | **8–13%** |

\* Ct values determined by clinical RT-qPCR from nucleic acid purified using the Hamilton Nimbus, as described in Methods section
† Based on validation data presented in this study
\*\*Lower sensitivity based on single reaction per sample, higher sensitivity based on duplicate reactions per sample

## Data Availability

Not applicable

